# Improving the representativeness of UK’s national COVID-19 Infection Survey through spatio-temporal regression and post-stratification

**DOI:** 10.1101/2023.02.26.23286474

**Authors:** Koen B. Pouwels, David W. Eyre, Thomas House, Ben Aspey, Philippa C. Matthews, Nicole Stoesser, John N. Newton, Ian Diamond, Ruth Studley, Nick G. H.Taylor, John I. Bell, Jeremy Farrar, Jaison Kolenchery, Brian D. Marsden, Sarah Hoosdally, E. Yvonne Jones, David I. Stuart, Derrick W. Crook, Tim E. A. Peto, A Sarah Walker, the COVID-19 Infection Survey Team

## Abstract

Population-representative estimates of SARS-CoV-2 infection prevalence and antibody levels in specific geographic areas at different time points are needed to optimise policy responses. However, even population-wide surveys are potentially impacted by biases arising from differences in participation rates across key groups. Here, we use spatio-temporal regression and post-stratification models to UK’s national COVID-19 Infection Survey (CIS) to obtain representative estimates of PCR positivity (6,496,052 tests) and antibody prevalence (1,941,333 tests) for different regions, ages and ethnicities (7-December-2020 to 4-May-2022). Not accounting for vaccination status through post-stratification led to small underestimation of PCR positivity, but more substantial overestimations of antibody levels in the population (up to 21%), particularly in groups with low vaccine uptake in the general population. There was marked variation in the relative contribution of different areas and age-groups to each wave. Future analyses of infectious disease surveys should take into account major drivers of outcomes of interest that may also influence participation, with vaccination being an important factor to consider.

## Introduction

The Covid-19 pandemic has a devastating impact on morbidity, mortality and economies around the world. As of 30 September 2022, there have been over 600 million confirmed Covid-19 cases, including over 6.5 million deaths according to the World Health Organization.^1^ These numbers substantially underestimate the true number of cases due to the lack of systematic testing in most countries. The United Kingdom (UK) has been a noticeable exception in terms of SARS-CoV-2 surveillance, recognising early on the value of investment in large population-based studies that follow a random sample of the population longitudinally with testing performed at fixed intervals independent of symptoms. This approach provides much more reliable estimates of levels and trajectories of the prevalence of SARS-CoV-2 infections and antibody levels than solely having to rely on national testing systems.^2^ In most countries, only people with specific symptoms, or those with contacts with known cases, are eligible for testing in systems set up by governments. However, a substantial proportion of individuals do not report any symptoms around their positive SARS-CoV-2 PCR test,^3^ and testing capacity, testing strategies and the probability that a symptomatic individual decides to get tested varies by time, socio-demographic factors and location.^2,4^ This complicates interpretation of such data sources that are not designed to provide representative estimates of prevalence or incidence of SARS-CoV-2 positive individuals. To inform decisions around implementation or (dis)continuation of (local) mitigation measures, policy makers ideally would have population-representative estimates of how many people are infected with SARS-CoV-2 in small areas at different time points. Similarly, it is important to track how SARS-CoV-2 antibody levels change over time to enable the likely levels of protection at the national and more granular regional levels to be accounted for when making vaccination and other mitigation policy decisions.

Here, we use data from the UK’s national COVID-19 Infection Survey (CIS) to demonstrate how a spatio-temporal regression and post-stratification modelling approach can be used to obtain representative temporal estimates of the swab positivity and antibody prevalence at the national and sub-regional level, and for different ages and ethnicities.^2,5-7^The UK’s CIS depends on voluntary participation. Therefore, despite invitations being sent to randomly selected addresses and monetary compensation for participation, it is possible that it is not optimally representative of the whole population. Here we explore to what extent accounting for vaccination status in the sample compared to the general population improves estimates of swab and antibody positivity, recognising the possibility that individuals who are more likely to get vaccinated may also be more likely to participate in infectious disease surveys upon invitation. In addition, we evaluate to what extent there is variation in trends within the nine regions of England and whether those areas that frequently have high swab positivity are more deprived and more urban.

## Results

Between Monday 7 December 2020 and Wednesday 4 May 2022, 6,496,052 PCR test results, taken following an external assessment schedule without knowledge of symptom status from participants in England, were available for analyses. Of these tests, 120,436 (1.9%) were positive. During the same period 1,941,333 blood samples from participants in England were tested for SARS-CoV-2 anti-spike IgG antibody levels, of which 1,738,778 (90%), 1,360,860 (70%), and 748,659 (39%) tests were above 23, 100, and 447 binding antibody units (BAU) per millilitre thresholds, respectively. The 23 BAU/ml threshold corresponds to the optimal diagnostic level to identify previous infection prior to vaccination, the 100 BAU/ml threshold to the antibody level estimated to confer 67% protection against Delta infection, and 447 BAU/ml threshold is the upper limit of quantification of the assay at its original 1:50 dilution, meaning this is the highest level that can be compared over time.^8^

### Swab positivity

There was marked variation in PCR positivity over time, with the estimated prevalence ranging from 0.09% in week 18 of 2021 to 7.35% in week 12 of 2022 in England. While the South-West region had lower PCR positivity for most of the study period, its Omicron BA.2 peak was more pronounced than in the London region that generally experienced higher prevalence of most variant waves, with a particularly high Omicron BA.1 peak (Fig. 1, Table S1). Similarly, while individuals of black ethnicity experienced a large BA.1 peak compared to other ethnicities, they experienced a less pronounced BA.2 peak than other ethnicities (Fig. S1). PCR positivity varied markedly between waves for different age-groups (Fig. S2). For example, adolescents aged 12-15 had by far the highest PCR positivity peak during the Delta wave, while their PCR positivity rates were consistently lower than for other age categories during the Omicron BA.2 wave.

**Fig 1:**
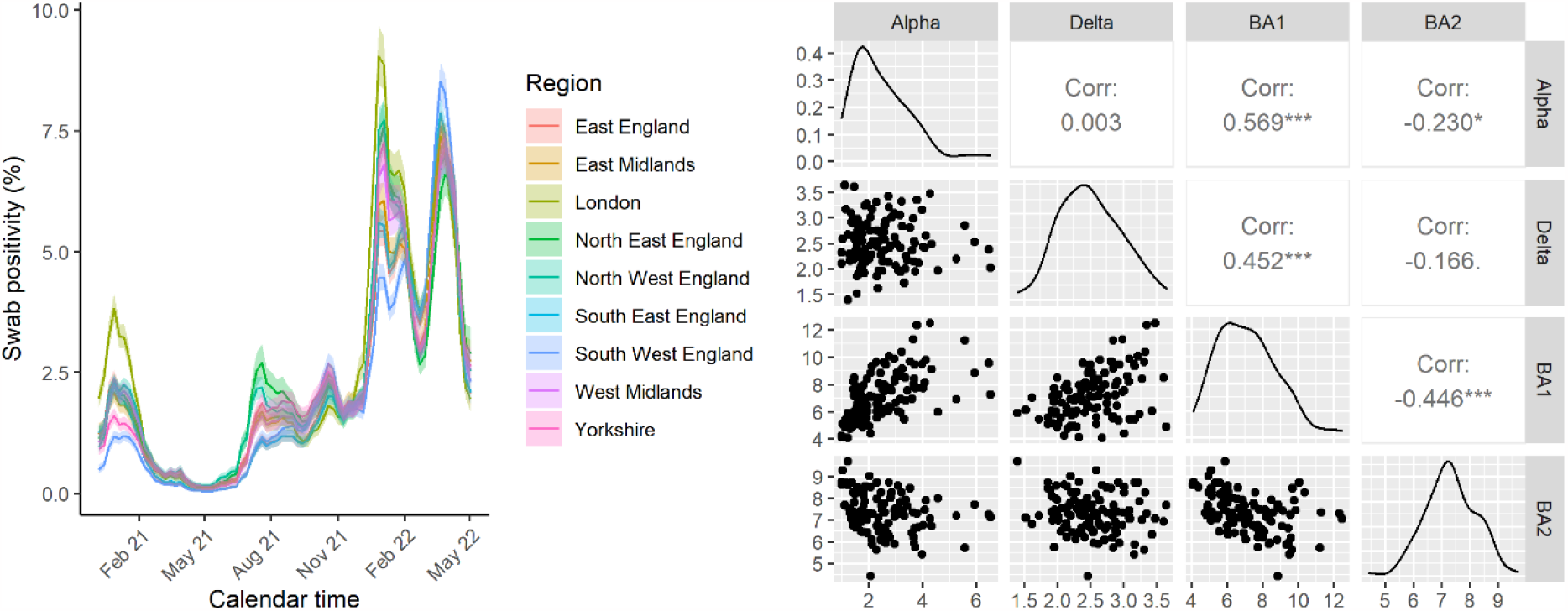
Post-stratified estimate of swab PCR positivity by region over time. Estimates are post-stratified for age, sex, CIS area (116 sub-regions within the 9 administrative regions shown), ethnicity and vaccination status. The right panel shows the correlations between the peaks of the Alpha, Delta, Omicron BA1, and Omicron BA2 among CIS areas. Correlations at the 9 administrative regions were qualitatively similar: Alpha-Delta 0.021; Alpha-BA1: 0.680; Alpha BA2: -0.302; Delta-BA1: 0.408; Delta-BA2: -0.117; BA1-BA2:-0.685.

Study participants were more likely to be vaccinated than the overall population (e.g. 93% in the survey vs 75% based on the admin data by May 2022, Fig. 2). While survey participants were more likely to be vaccinated than expected based on the national administrative data on vaccination uptake, post-stratifying for vaccination status had only small effects on estimated levels of PCR positivity (Fig. 2). When not accounting for over-representation of vaccinated individuals in the survey, the largest underestimation of positivity across England overall was 0.38% (6.30% vs 6.69%). When focusing on subregional estimates of swab positivity, dividing the nine regions of England into 116 sub-regions (CIS areas), the largest differences were observed for areas within London, with a maximum difference in point estimates of 1.28% (10.82% vs 12.10%) in Lambeth during the first week of 2022.

**Fig. 2:**
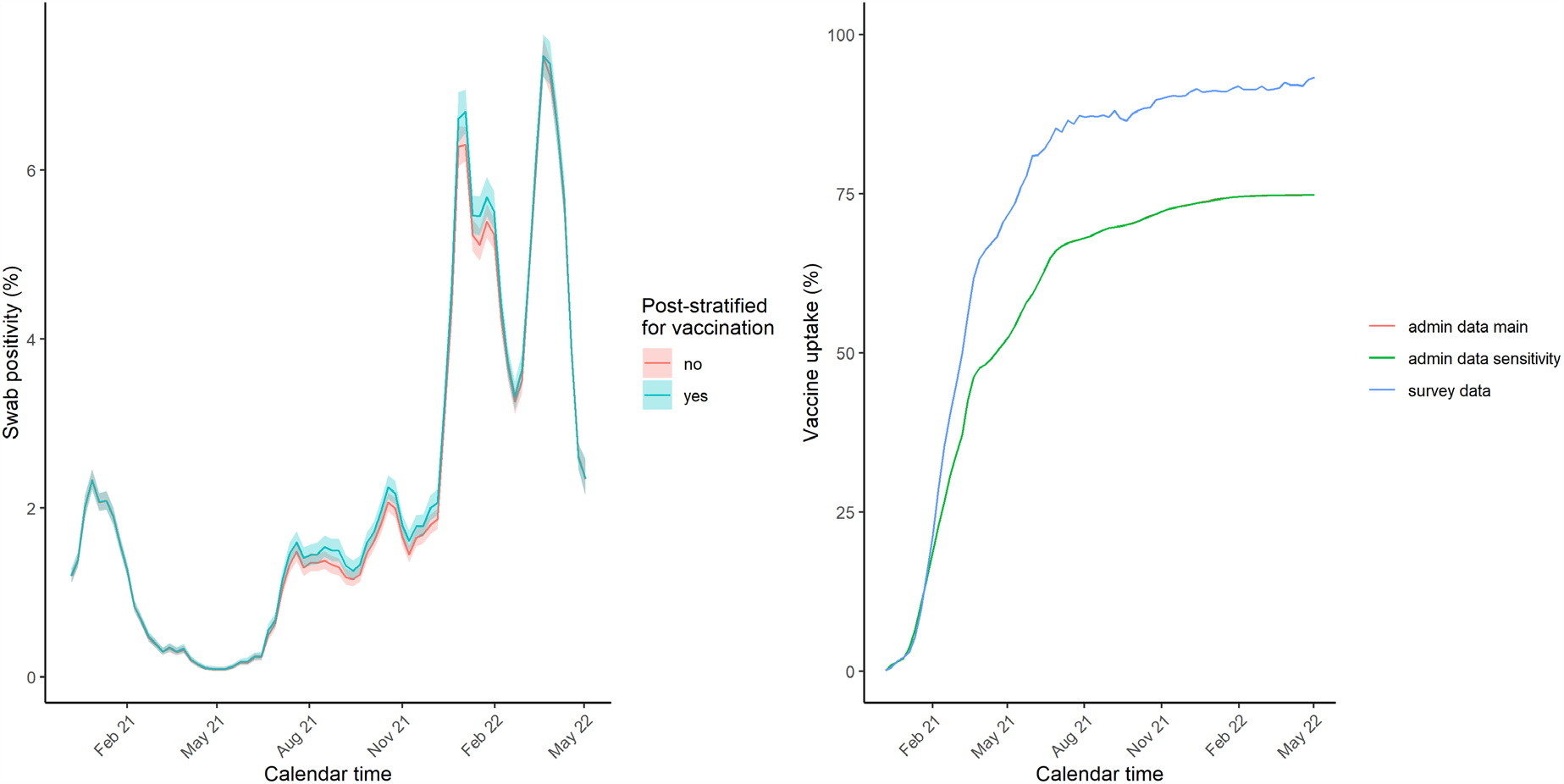
Impact of post-stratifying for vaccination status (yes/no and interaction with time) on swab PCR positivity and crude cumulative vaccination uptake based on survey participants and admin data. The red and green line almost perfectly overlap reflecting the fact that the assumptions we made about vaccination uptake among children too young to be present in the 2011 Census are of negligible importance given that during the study period very few children aged 9-11 years of age – the ages from which we extrapolated to younger children assuming I) no vaccinations under the age of 5 and half the coverage of that observed among 9-11 years old children for those aged 5-8 years of age (main analysis), or II) no vaccinations under the age of 9 (sensitivity analysis).

There was large variability over time in which areas of England had the highest PCR positivity, but some areas also consistently had higher or lower PCR positivity (Fig. 3, Fig S3). Using the spatiotemporal regression and post-stratification model that accounted for vaccination status, we evaluated whether certain areas consistently had a high probability (≥80%) of being ranked among the top 10 areas in terms of the highest weekly PCR positivity (Fig. 4). Three areas out of a total of 116 areas (2.6%) – Kirklees, Rochdale, and Nottingham – had a high probability of being ranked in the top 10 areas of highest swab positivity over more than 25% of the study period (>18 out of 74 weeks), while many other areas were never ranked in the top 10 (Table 1 and S2). A linear regression with area-specific levels of deprivation (0.25, 95% CI 0.16-0.34 decrease per 1 unit increase in deprivation ranking; t statistic -5.49) and the percentage of the area considered rural (0.46, 95% CI 0.34-0.59 decrease per 1% increase rurality; t statistic -7.25) as the only covariates explained 56% of the variance in median PCR positivity ranking of the areas, indicating that less deprived areas and more rural areas were likely to have lower PCR positivity compared to more deprived and urban areas.

**Table 1.** Ranking of CIS areas in terms of swab positivity.

| **CIS area** | **Region** | **Number of times with a >=80% probability of being in top 10 areas with high swab positivity (N=74 weeks)** | **Average deprivation score rank of areas within the CIS area(1 most deprived)** | **% of total population in the CIS area that lives in most deprived (10%) areas nationally** |
| --- | --- | --- | --- | --- |
| Kirklees | Yorkshire | 28 | 47 | 12% |
| Nottingham | East Midlands | 22 | 5 | 31% |
| Rochdale | East Midlands | 21 | 8 | 30% |
| Burnley, Hyndburn, Pendle, Rossendale, Bury | North West | 15 | 24 | 22% |
| Barking and Dagenham | London | 14 | 11 | 4% |
| Blackburn with Darwen, Chorley, Bolton | North West | 13 | 23 | 23% |
| Newham | London | 13 | 22 | 2% |
| Manchester | North West | 11 | 2 | 43% |
| Tower Hamlets | London | 10 | 27 | 1% |

**Fig. 3:**
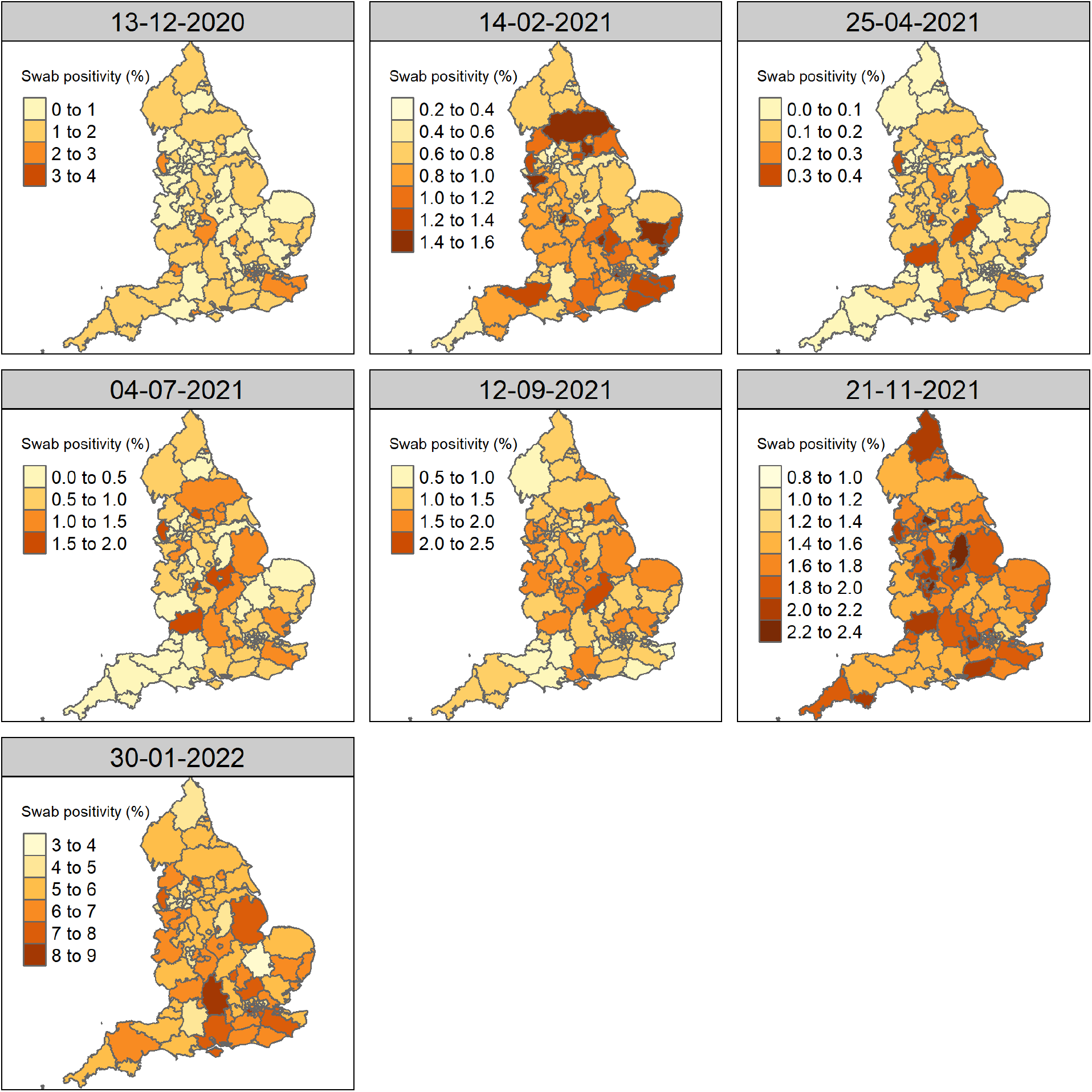
Post-stratified estimate of swab PCR positivity by CIS area over time. Estimates are post-stratified for age, sex, ethnicity and vaccination status.

**Fig 4:**
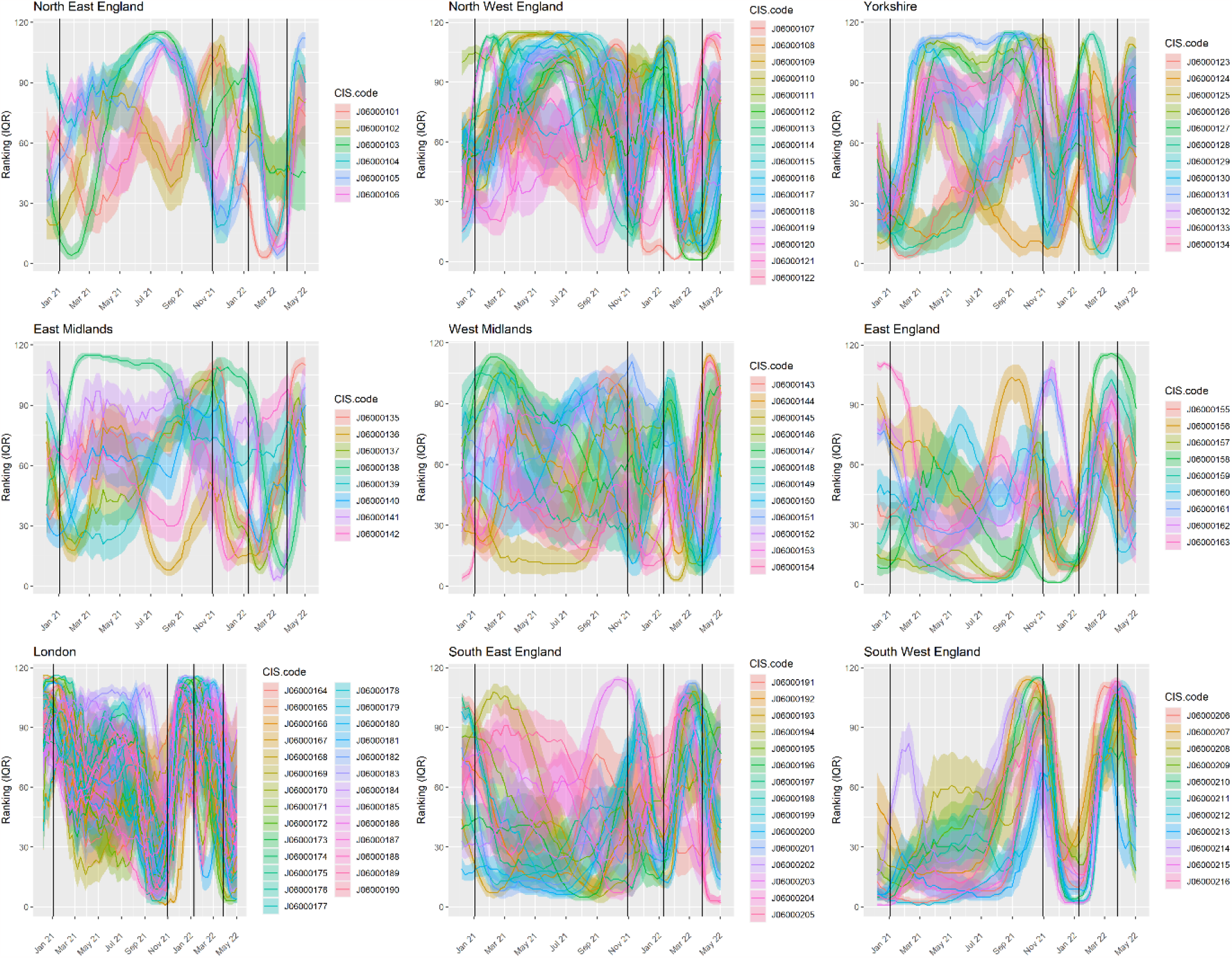
Median ranking of CIS areas in terms of swab positivity. A low ranking corresponds to a CIS area having lower swab positivity estimates at that point in time compared to other CIS areas in the country. Shaded areas represent the interquartile range of the ranking of the CIS areas. Vertical black lines indicate from left to right the peaks (at the national level) of the Alpha, Delta, Omicron BA1, and Omicron BA2 wave, respectively.

### Antibody prevalence

We primarily estimated antibody prevalence at a threshold previously estimated to be associated with 67% protection against new infection with the Delta variant (100 BAU/ml) (results similar at other thresholds, table S1).^8^ Antibody positivity at this threshold during the first week of the vaccination campaign in England, which started on 8 December 2020, was only 5.61% (95% CI 4.88-6.43%). The percentage of individuals having antibodies levels ≥100 BAU/ml increased over time, with steeper increases coinciding with increases in second and third vaccinations (Fig. 5).

**Fig. 5:**
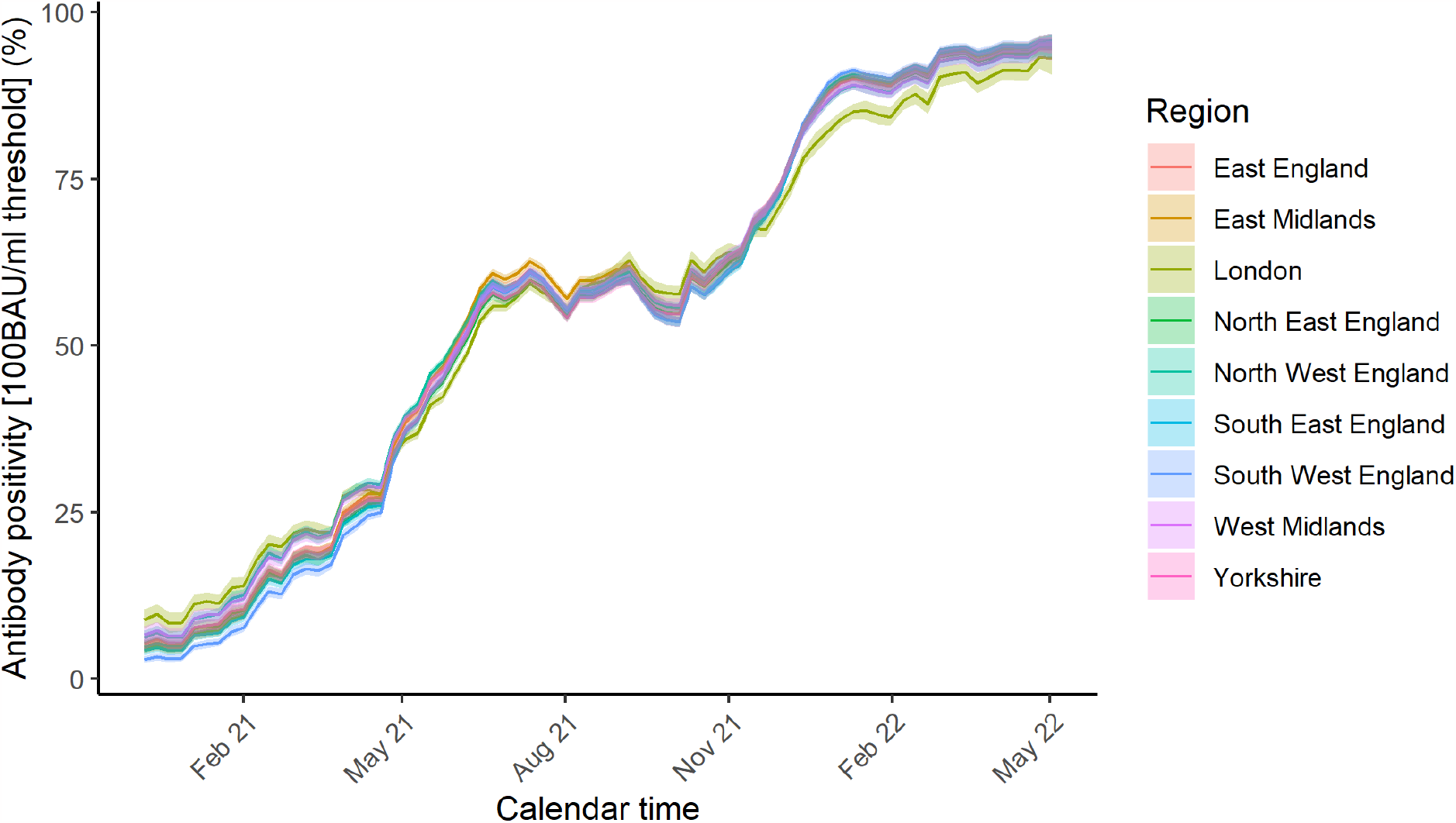
Post-stratified estimate of antibody positivity at the 100 BAU/ml threshold by region over time. Estimates are post-stratified for age, sex, CIS area, ethnicity and vaccination status.

Post-stratifying for vaccination status had negligible effects at the start of the vaccination campaign, but over time, when an increasing proportion of individuals had antibody levels above 100 BAU/ml due to vaccination rather than infection, the effect of accounting for over-representation of vaccinated individuals in the survey became more marked (Fig. 6). Not accounting for vaccination status led to point estimates for antibody positivity (≥100 BAU/ml) that were on average 4.8% higher than when taking this into account, with the largest difference (21%; 77% vs 54%) observed in Newham (London region) during the third week of July 2021. When using the spatiotemporal models to post-stratify and summarise positivity estimates by different population characteristics, differences between the models with and without vaccination status were most marked in areas and population groups with lower vaccine uptake according to the administrative data, e.g., for non-white ethnicities with the largest differences observed for black ethnicity (Fig. S4), and certain age-categories with the largest differences in estimates occurring among those aged 25-34 years old (Fig. S4).

**Fig 6:**
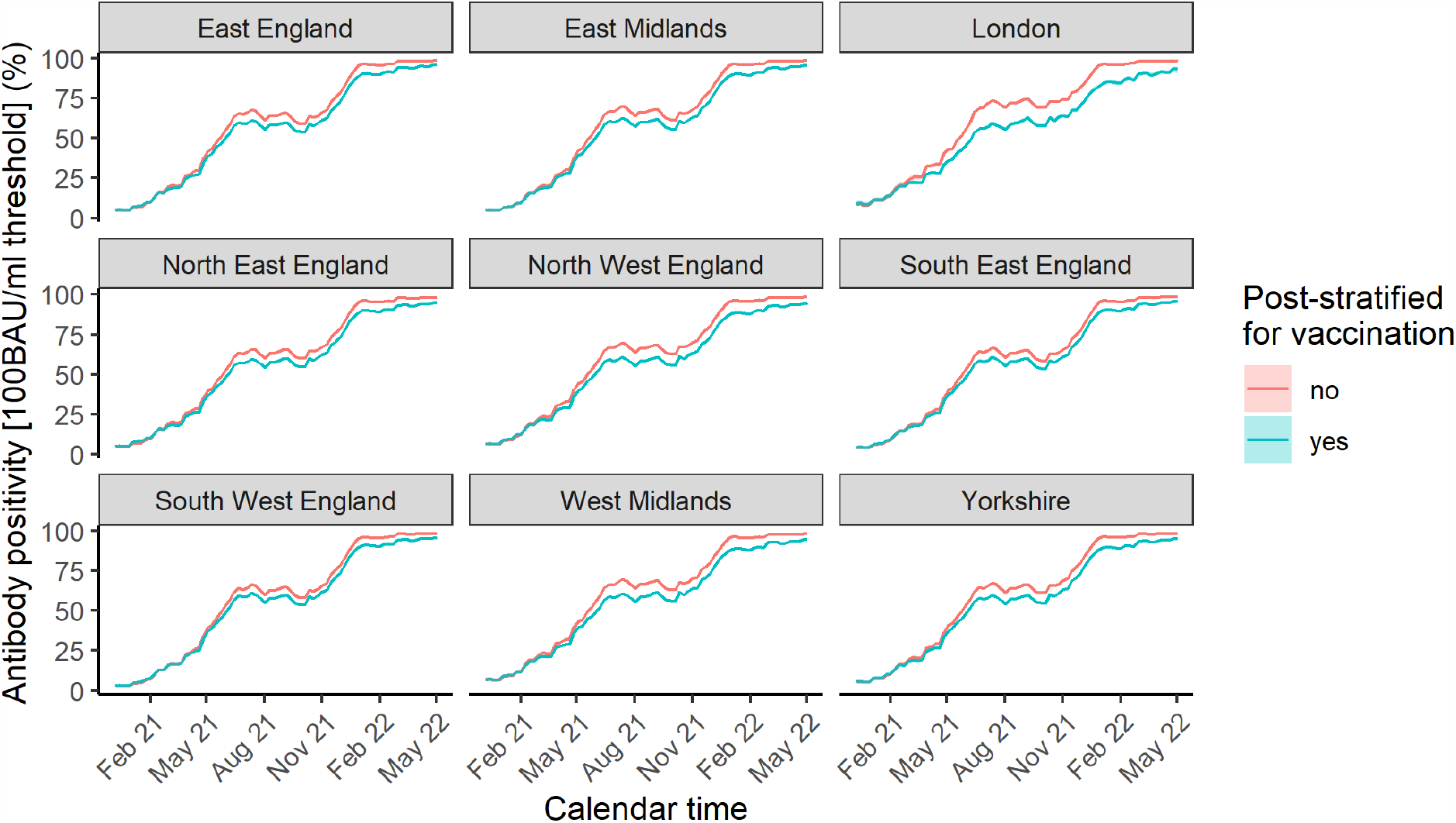
Impact of post-stratifying for vaccination status (yes/no and interaction with time) on estimated antibody positivity at the 100 BAU/ml threshold by region over time. Estimates are post-stratified for age, sex, CIS area, ethnicity and vaccination status.

Subsequently, using models that post-stratified for vaccination status, we evaluated whether certain areas consistently had a high probability (≥80%) of being ranked among the top 10 areas in terms of the highest antibody positivity at the 100 BAU/ml threshold. At the start of the vaccination campaign this will have been almost exclusively driven by antibody responses to previous infection, but over the course of the campaign, antibody levels above the threshold would be increasingly due to vaccinations alone or a combination of vaccinations and previous infections. Consistent with these different drivers, several areas in London that initially had relatively high antibody levels due to higher rates of previous infections were subsequently among the areas with the lowest percentage with antibody levels above the threshold due to lower vaccination uptake (Fig. 7).

**Fig. 7:**
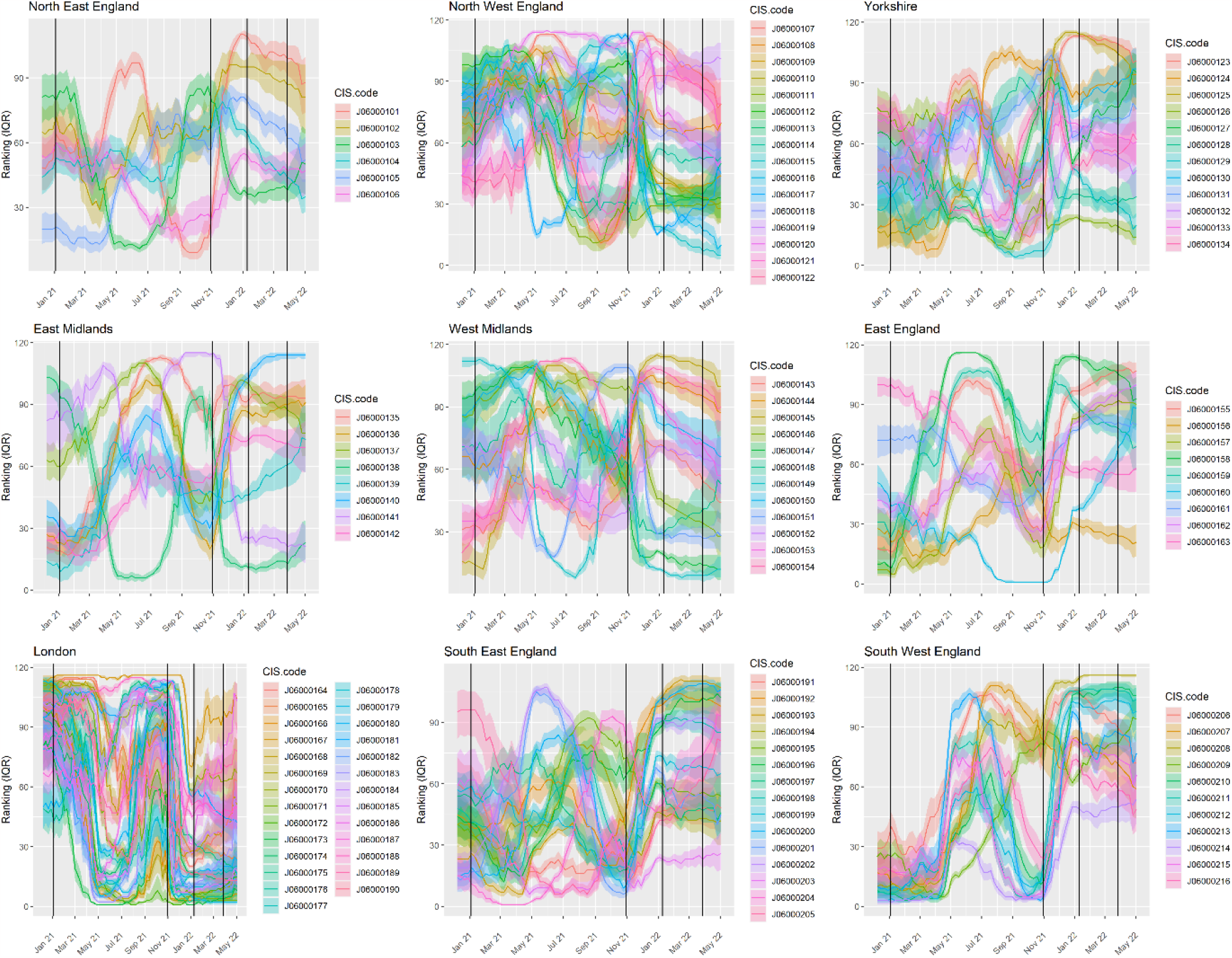
Median ranking of CIS areas in terms of antibody positivity at the 100 BAU/ml threshold. A low ranking corresponds to a CIS area having lower antibody positivity estimates at that point in time compared to other CIS areas in the country. Shaded areas represent the interquartile range of the ranking of the CIS areas. Vertical black lines indicate from left to right the peaks (at the national level) of the Alpha, Delta, Omicron BA1, and Omicron BA2 wave, respectively.

Area-specific levels of deprivation and the percentage of the area considered rural explained only 10% of the variation in median ranking for antibody prevalence when considering the entire study period from the start of the vaccination campaign to May 2022, when Omicron variants dominated. To assess whether this lower percentage of the variance being explained may be due to deprived and urban areas being associated with higher infection rates but lower vaccination uptake - effects that would cancel out to a certain extent - we considered distinct time periods separately. Before March 2021, when <2% of the population had received their second vaccination, this simple model with two covariates explained 43% of the variation in the median ranking for antibody prevalence (0.14, 95% CI -0.02-0.29 decrease per 1 unit increase in deprivation ranking; t statistic -1.78; 0.80, 95% CI 0.59-1.01 decrease per 1% increase rurality; t statistic -7.43). In contrast, from 3 January 2022 onwards - when a large proportion of the population had received at least 3 vaccinations - the same model explained 67% of the variance, but with lower levels of deprivation (higher deprivation ranking) and a higher percentage of the area being considered rural being associated with relatively high antibody prevalence (0.43, 95% CI 0.31-0.55 increase per 1 unit increase in deprivation ranking; t statistic 7.31; 0.73, 95% CI 0.57-0.90 increase per 1% increase rurality; t statistic -8.81).

## Discussion

Using data from one of the largest SARS-CoV-2 community surveillance studies in the world, that randomly invites individuals from private households to obtain representative estimates of SARS-CoV-2 PCR positivity and antibody levels, we found that those who agree to participate in the survey are more likely to be vaccinated than the overall population. Not accounting for vaccination status, as done throughout the pandemic, resulted in a small underestimation of PCR positivity but a more substantial overestimation of the percentage of the population having antibody levels at least as high as the threshold previously estimated to be associated with 67% protection against infection with the Delta variant.^8^ While estimates were, as expected, similar with and without accounting for vaccination status at the start of the vaccination campaign, and 4.8% when taking the average difference for England over the entire period, the maximum difference in antibody prevalence at the aforementioned antibody threshold was 21%. Such large differences, observed in an area with one of the lowest percentages of White British populations and one of the highest poverty rates in England, could lead to underinvestment in interventions that could help increase the percentage of the local population with sufficiently high antibody levels to be protected against new SARS-CoV-2 infections, potentially contributing to severe acute disease or long COVID-19 in the population.

Importantly, these findings suggest that when performing infectious disease surveillance for pathogens for which vaccines are available and commonly used, accounting for vaccination status should be recommended, as despite randomly inviting individuals and compensating them for participation, those that decide to participate in the survey may be more likely to be vaccinated than those that decide to decline the invitation. Accounting for vaccination status through post-stratification could have a particularly large impact among groups of the population with lower vaccination rates, as here also observed for non-white ethnicities, younger age groups, and those living in urban and more deprived areas.^9^ These findings support the suggestion that even large studies that aim to randomly select individuals from the target population have to carefully consider how to prevent and minimise selection bias and account for vaccination status when using carefully designed surveys for infectious diseases.^10^ For example, the Census Household Pulse conducted by the US Census Bureau and eleven statistical government partners, substantially overestimated uptake of the first dose vaccination by 14% in May 2021 despite accounting for age, gender, education, and ethnicity.^10^ In comparison, on 23 May 2021, crude estimates of uptake of the first dose vaccination were approximately 20% higher in the CIS compared to estimates based on the administrative data.

Areas with higher levels of deprivation and more urban areas more often had a higher PCR positivity than other areas, explaining a large percentage of the observed variation in median ranking of PCR positivity. The same was true for antibody positivity prior to vaccination becoming widely available, and after Omicron variants dominated. These associations occurred only when splitting the study into separate periods because high levels of deprivation and urban areas had higher infection rates but lower vaccination uptake, resulting in effects cancelling each other out to a certain degree when focusing on antibody prevalence across the entire period.

Deprivation and percentage of the area that was rural were not included in the main PCR and antibody positivity models to avoid artificial associations from using the same variables to predict the prevalence in the target population and subsequently in a separate model to assess the relationship between the same variable and post-stratified prevalence. Furthermore, both variables are only readily available as area-based markers and partly already captured by post-stratifying to relatively small CIS areas within England. Ideally one would be able to account for individual-based socio-economic status related variables, such as educational qualifications and household assets, among survey participants and the general population (the target population) through linkage to the 2021 Census in England.^11^ However, this was not available for research at the time of the current analysis.

The validity of the post-stratification relies on the absence of model misspecification, e.g. not missing an important variable that both influences the decision to participate in the survey upon invitation and the outcomes considered here. Variables that are associated with the decision not to participate in surveys like this despite monetary compensation, which may increase the probability that those of lower socio-economic status participate,^12^ may also be associated with behaviour that increases the risk of acquiring an infection with SARS-CoV-2. This may have led to underestimating swab positivity, and hence antibody levels mediated through swab positivity, while characteristics that mainly affect outcomes through vaccination uptake are less relevant as we took into account vaccination status.

Model complexity meant that we could only allow for whether participants were vaccinated as a binary variable (yes/no) interacting with time. More complicated models with the number of vaccinations failed to converge without errors due to very high/low positivity rates during large parts of the study period. Therefore, we implicitly assume that the survey - after conditioning on CIS area, age, sex, ethnicity, and time – may not be representative in terms of whether individuals decide to get any COVID-19 vaccine, but that - after conditioning on the same variables - vaccinated individuals in the survey are representative of vaccinated individuals in the target population.

Whether the post-stratification used effectively removes any bias due to non-response is also dependent on how accurate the information on conditional distributions of variables are in the target population. While the number of vaccinated individuals are well-recorded through the National Immunisation Management Service (NIMS) system, there is more uncertainty about the number of individuals in each subgroup of the population that did not get vaccinated as it is not exactly known how many people live in England. For the current study, we restricted to individuals that could be linked to 2011 Census to ensure a well-defined denominator. Consequently we had to make assumptions about the uptake of individuals aged 2-8 years old. However, the PCR positivity estimates were for the 2-11 years old were not sensitive to the assumptions made, and we only evaluated antibody levels for those aged 16years and above. The estimated number of unvaccinated individuals in each subgroup relies on the assumption that more recent migrants have similar rates of uptake as individuals of the same age, sex, ethnicity, and location as those that were already present in England during the 2011 Census. If this assumption does not hold, the administrative data on vaccination uptake may also be not completely accurate. As information from the 2021 Census is released, these may be updated.

In conclusion, we have shown that not accounting for vaccination status overestimates the percentage of people that still have sufficient antibody levels to be protected against new infection, potentially affecting decision making. Future analysis of the CIS and other surveys should, whenever possible, account for major drivers of the outcome of interest that are also likely associated with non-response to invitations to participate, with vaccination being particularly important when looking at infectious disease outcomes such as SARS-CoV-2 antibody levels. Using a spatiotemporal model that accounts for vaccination status, we have shown substantial variation between and within regions of England over the course of the pandemic and identified areas that had a high probability of having a higher SARS-CoV-2 prevalence than other areas throughout a large part of the pandemic. A large part of the variation in the ranking of small-areas in terms of their SARS-CoV-2 prevalence could be explained by the degree of urbanicity and deprivation, highlighting the inequality in risk of SARS-CoV-2 infections and its subsequent consequences.

## Methods

### Study participants

Data were obtained on all SARS-CoV-2 RT-PCR test results between 8 December 2020 and 04 May 2022 from nose and throat swabs taken from individuals participating in the Office for National Statistics (ONS) CIS (ISRCTN21086382, https://www.ndm.ox.ac.uk/covid-19/covid-19-infection-survey/protocol-and-information-sheets) living in private households in England. We restricted the current analyses to England, as detailed administrative data on vaccination uptake from which to construct post-stratification tables (see below) were only available for England. The survey randomly selects private households on an ongoing basis from address lists and previous surveys to provide a representative UK sample. Details on the sampling design are provided elsewhere.^2^ Following verbal agreement to participate, a study worker visited each household to take written informed consent. This consent was obtained from parents/carers for those 2-15 years, while those 10-15 years also provided written assent. Children aged <2 years were not eligible for inclusion into the study.

Individuals were also asked about demographics and vaccination uptake (https://www.ndm.ox.ac.uk/covid-19/covid-19-infection-survey/case-record-forms). At the first visit, participants were asked for (optional) consent for follow-up visits every week for the next month, then monthly for 12 months from enrolment. Initially, in a random 10-20% of households, those 16 years or older were invited to provide blood monthly for assays of anti-trimeric spike protein IgG using an immunoassay developed by the University of Oxford.^13,14^ Household members of participants who tested positive were also invited to provide blood monthly for follow-up visits. These participants were excluded from the analysis to avoid overestimation of antibody levels. From April 2021, additional participants were invited to provide blood samples monthly to assess vaccine responses, based on a combination of random selection and prioritisation of those in the study for the longest period (independent of test results).^8,15,16^

The study received ethical approval from the South Central Berkshire B Research Ethics Committee (20/SC/0195).

### Swab positivity

Nose and throat self-swabs were couriered directly to the UK’s national Lighthouse laboratories (National Biocentre in Milton Keynes and Glasgow) where samples were tested as part of the national testing programme. Identical methodology was used to test for the presence of SARS-CoV-2 genes for nucleocapsid protein (N), spike protein (S), and ORF1ab using RT-PCR.^2^ We used the TaqPath RT-PCR COVID-19 kit (Thermo Fisher Scientific, Waltham, MA, USA), which was analysed using UgenTec Fast Finder 3.300.5 (TagMan 2019-nCoV assay kit V2 UK NHS ABI 7500 v2.1; UgenTec, Hasselt, Belgium). The assay plugin contains an assay-specific algorithm and decision mechanism that allows conversion of the qualitative amplification assay PCR raw data from the ABI 7500 Fast into test results with little manual intervention. Samples are called positive in the presence of at least one gene (N, ORF1ab, or both) but could be accompanied by the gene for S protein (ie, one, two, or three gene positives). The gene for S protein is not considered a reliable single gene positive.^2^

### Antibody prevalence

Blood samples were couriered to the clinical biochemistry and microbiology laboratories at the John Radcliffe Hospital in Oxford to test for the presence of antibodies using the Oxford immunoassay.^2,14^ Normalised results are reported in ng ml^−1^ of mAb45 monoclonal antibody equivalents. Before 26 February 2021, the assay used fluorescence detection as described previously, with a positivity threshold of 8 million units validated on banks of known SARS-CoV-2 positive and SARS-CoV-2 negative samples.^8,14^

After this, it used a commercialized CE-marked version of the assay, the OmniPATH 384 Combi SARS-CoV-2 IgG ELISA (Thermo Fisher Scientific), with the same antigen and colorimetric detection. mAb45 is the manufacturer-provided monoclonal antibody calibrant for this quantitative assay. The fluorometrically determined values were converted into arbitrary units using the following conversion formula:

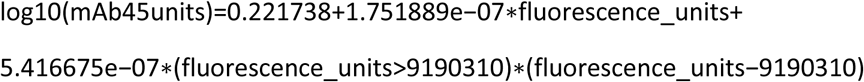

The results of the OmniPATH assay were converted into WHO international international units (BAU ml^−1^) using the following formula:

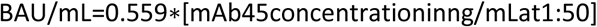

We used ≥23 BAU/ml as the threshold for determining IgG positivity (corresponding to the 8 million units with fluorescence detection). The upper limit of quantification of the assay is 447 BAU/ml at the standard 1:50 dilution.^8^ From 28 January 2022, samples were tested at 1:400 dilution, and from 29 April 2022 at 1:1600 dilution, with those below the lower limit of quantification at these dilutions retested at the original 1:50 dilution.

### Vaccination uptake

Participants were asked about their vaccination status at study visits, including information about the type of vaccine, the number of doses received to date, and the date of the most recent vaccination. For England, linked administrative vaccine uptake data is also available from the National Immunisation Management Service (NIMS), which records details of all COVID-19 vaccinations provided by the National Health Service (NHS) in England. NIMS covers the entire population of England, but does not include vaccinations obtained abroad. The Office for National Statistics provided weekly estimates of the proportion of individuals in each subgroup of the population based on NIMS data linked to Census 2011 data. Therefore, only participants that were already living in England during the Census 2011 were used to inform conditional estimates of vaccine uptake, thereby implicitly assuming that vaccination uptake in each subgroup of the population is the same for individuals from that subgroup living in England as those from the same subgroup of the population that moved to the England more recently. These age-, sex-, ethnicity-, CIS area-, and time-specific estimates of vaccine uptake were subsequently applied to 2020 population estimates provided by the Office for National Statistics.

### Statistical analyses

Bayesian multilevel regression and poststratification is an increasingly used statistical technique to obtain representative estimates of prevalence or preferences at the national and smaller regional levels.^2,6,17-22^ This method has been found superior at both national and regional levels compared to traditional survey weighted and unweighted approaches in several empirical and simulation studies.^2,6,17-22^ By using random effects in the multilevel model, stable estimates can be obtained for sub-national areas from relative small samples or relatively rare outcomes.^2^ However, if there is an spatial underlying structure this needs to be captured by the multilevel regression and post-stratification (MPR) methodology to avoid biased estimates based on a model that assumes independent group-level errors. Gao et al. recently proposed a spatial MRP model using a Besag-York-Mollié (BYM2) specification for the regional effect.^6,23,24^ Using a simulation, they showed that when a spatial structure does exist, a spatial MRP with a BYM2 spatial prior for region improved MRP estimates through a reduction in absolute bias compared to using a default independent and identically distributed (IID) prior for the region effect. Importantly, when a spatial structure was not present in the underlying data, using a spatial MRP with a BYM2 spatial prior on region resulted in virtually identical posterior estimates as an MRP with an IID prior for region, suggesting that the BYM2 spatial prior does not force a spatial structure when it is not present.^6^

Here we extend the spatial MRP approach proposed by Gao et al. to a spatio-temporal context by adding a temporal component to the model.^5,6^ For the temporal components we used autoregressive or random walk processes with discrete time indices (weeks) to capture likely temporal effects in the MRP model. The choice of the type of directed conditional distribution for the time effect (random walk or autoregressive) type of space-time interaction (type I-IV) and inclusion of additional covariates is guided by comparing the Watanabe-Akaike information criterion (WAIC) of the models.^25,26^ A type I space-time interaction assumes no spatial and/or temporal structure on the interaction, a type II space-time interaction assumes that for the *i*th area the parameter vector has an autoregressive structure on the time component, which is independent from the ones of the other areas; a type III space-time interaction assumes that the parameters of the *t*th time point have a spatial structure independent from the other time points; while a type IV space-time interaction assumes that the temporal dependency structure for each area also depends on the temporal pattern of the neighbouring areas.^25^

The same set of covariates and interactions were considered for the MRP models for all outcomes (swab positivity and antibody prevalence at different thresholds (23, 100, and 477 BAU/ml)) and included: age (2-11, 2-15, 16-24, 25-34, 35-49, 50-59, 60-64, 65-69, 70-74, 75-79, 80+) (models for antibodies only included individuals aged 16 years or older given that blood samples were not taken in those aged <16y before November 2021); sex; ethnicity (Asian, Black, Mixed, White, Other); region (9 regions in England); CIS area (116 CIS areas, nested within regions in England); and two-way interactions of age, sex, and ethnicity with time and CIS or region area.

For each outcome, after running the final spatiotemporal binomial regression model, post-stratification was used to obtain representative estimates of the outcome prevalence in the target population. Post-stratification tables were based on the conditional distribution of age and sex by area from ONS. The conditional distribution of ethnicity by these categories were obtained from the ETHPOP database.^5,27^ Conditional distribution of vaccination uptake were obtained from the NIMS administrative data on vaccination uptake.

Using the population sizes of each poststratification cell of the target population, MRP adjusts for residual non-representative by post-stratifying by the percentage of each type in the actual overall population. If the model is correctly specified, unbiased estimates of prevalences at both national as well as sub-national and within categories can be obtained.

### Ranking of CIS areas in terms of prevalence

National testing data suggested that certain sub-regional areas in England had rather consistently higher prevalence of SARS-CoV-2 infections than the rest of the country, but it is not clear to what extent this is explained by testing behaviour or a true difference. Leveraging the fact that the applied models are Bayesian and the CIS is taking a random sample of the population, we evaluated the weekly probability that each CIS area is among the top 10 areas with the highest swab positivity prevalence.

Furthermore, we assessed to what extent the median ranking of CIS areas can be explained by area-specific levels of deprivation and degree of urbanicity using linear regression. Linear regression with deprivation and urban/rural classification (modelled as a continuous variable) as covariates was used.

## Data Availability

Data are still being collected for the COVID-19 Infection Survey. De-identified study data are available for access by accredited researchers in the ONS Secure Research Service (SRS) for accredited research purposes under part 5, chapter 5 of the Digital Economy Act 2017. Individuals can apply to be an accredited researcher using the short form on https://researchaccreditationservice.ons.gov.uk/ons/ONS_registration.ofml. Accreditation requires completion of a short free course on accessing the SRS. To request access to data in the SRS, researchers must submit a research project application for accreditation in the Research Accreditation Service (RAS). Research project applications are considered by the project team and the Research Accreditation Panel (RAP) established by the UK Statistics Authority at regular meetings. Project application example guidance and an exemplar of a research project application are available. A complete record of accredited researchers and their projects is published on the UK Statistics Authority website to ensure transparency of access to research data. For further information about accreditation, contact or visit the SRS website.

---

**Fig. S1.**
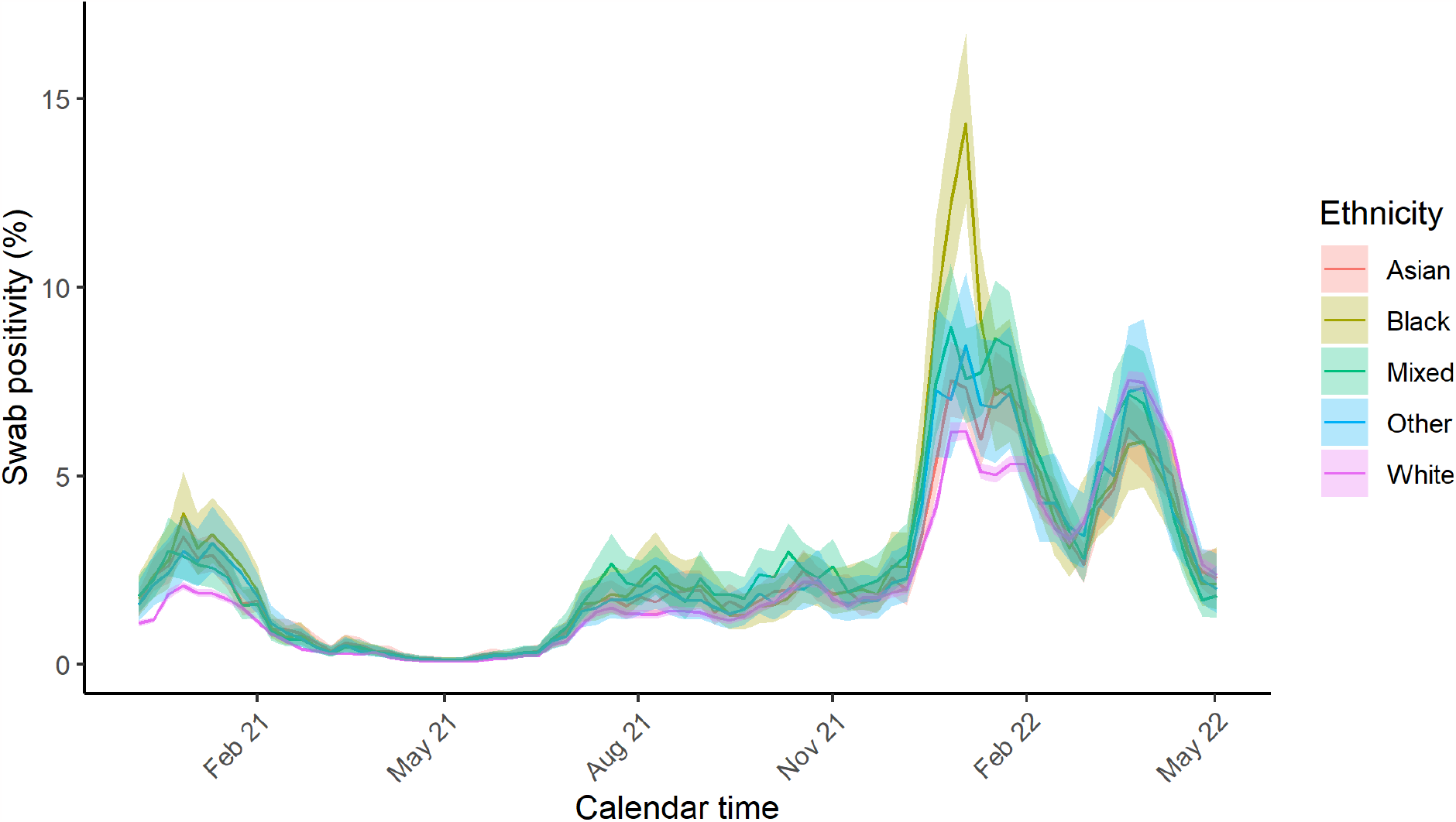
Post-stratified estimate of swab positivity by ethnicity over time. Estimates are post-stratified for age, sex, CIS area, ethnicity and vaccination status.

**Fig. S2.**
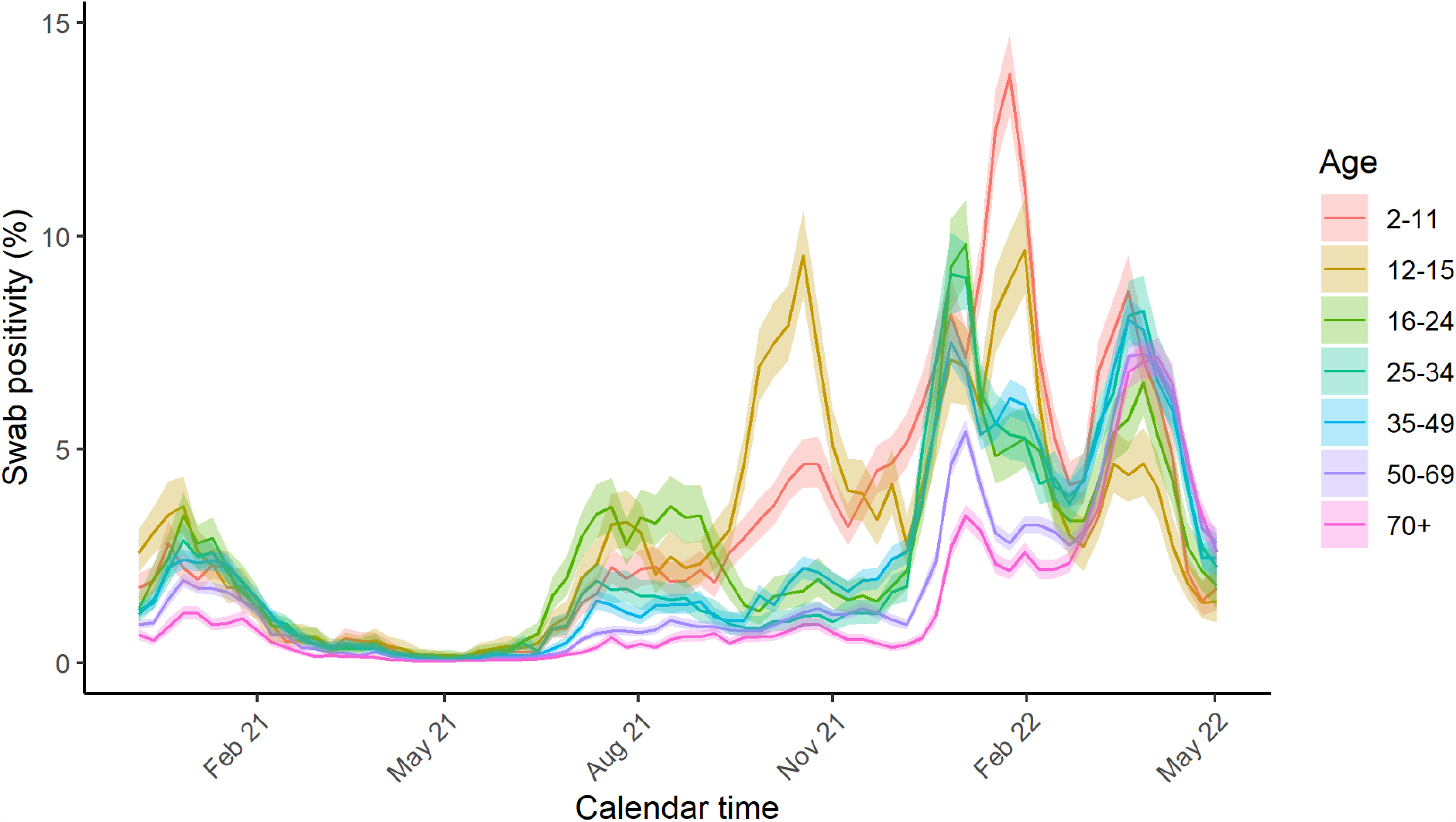
Post-stratified estimate of swab positivity by age-group over time. Estimates are post-stratified for age, sex, CIS area, ethnicity and vaccination status.

**Fig. S3:**
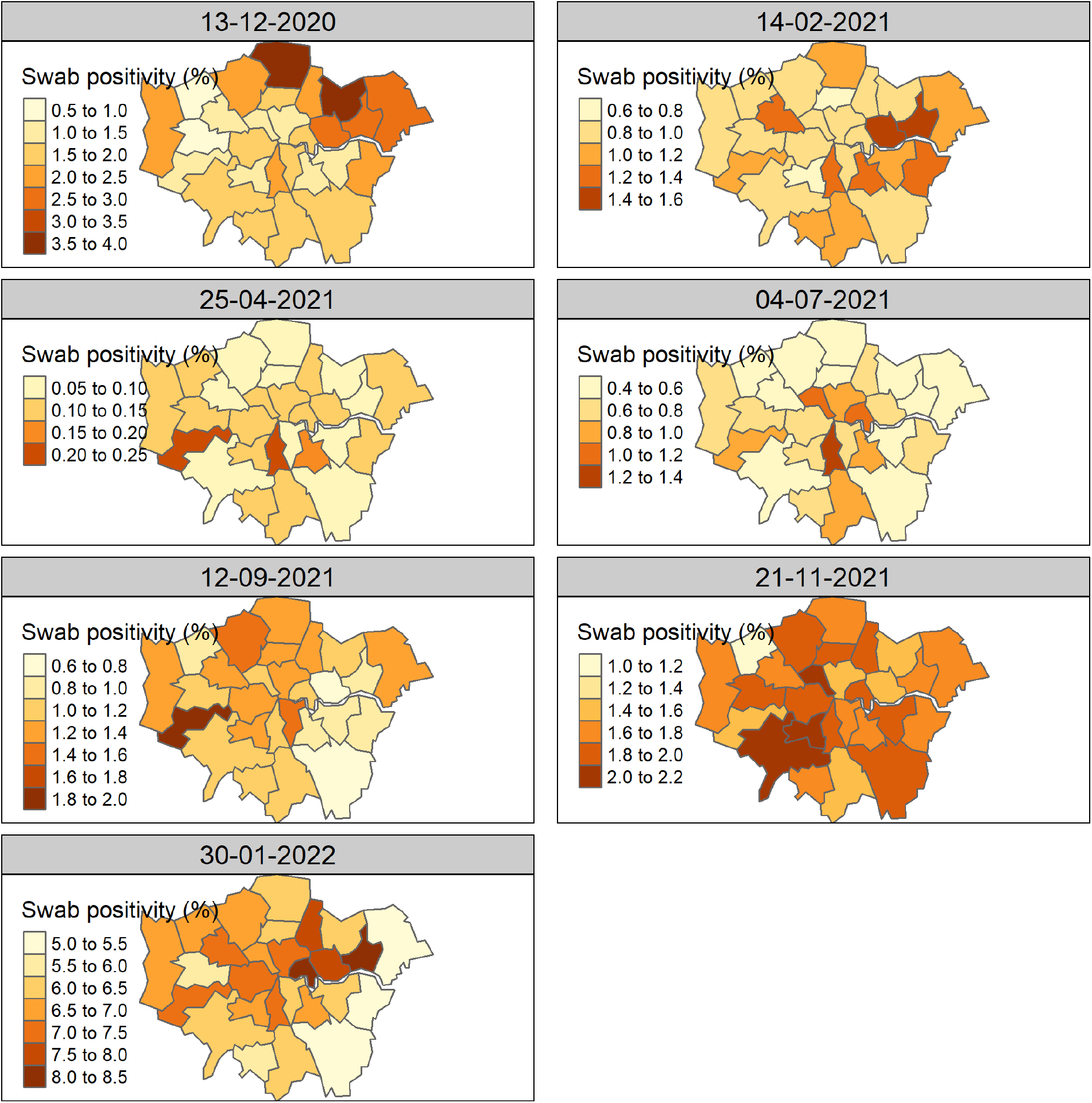
Post-stratified estimate of swab PCR positivity by CIS area in London over time. Estimates are post-stratified for age, sex, CIS area, ethnicity and vaccination status.

**Fig S4.**
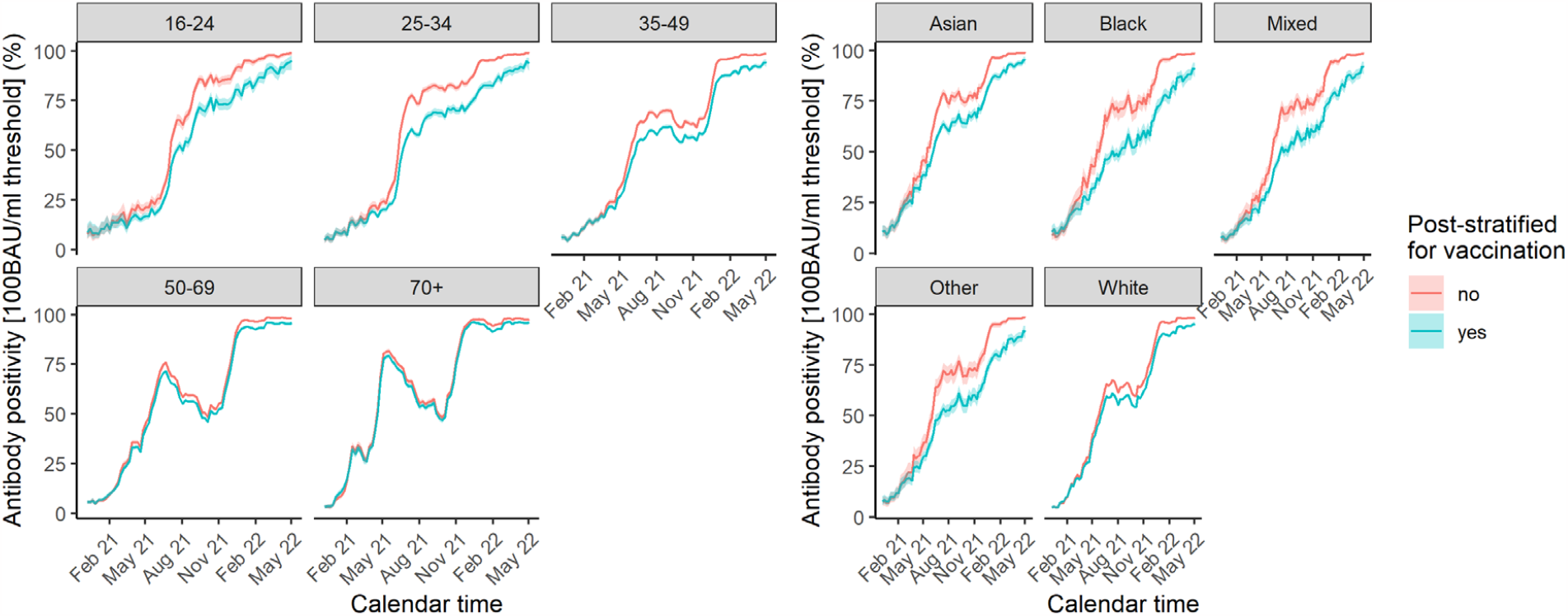
Impact of post-stratifying for vaccination status (yes/no and interaction with time) on estimated antibody positivity at the 100 BAU/ml threshold by age and ethnicity over time. Estimates are post-stratified for age, sex, CIS area, ethnicity and vaccination status.

